# Social relationships and depression during the COVID-19 lockdown: longitudinal analysis of the COVID-19 social study

**DOI:** 10.1101/2020.12.01.20241950

**Authors:** Andrew Sommerlad, Louise Marston, Jonathan Huntley, Gill Livingston, Gemma Lewis, Andrew Steptoe, Daisy Fancourt

## Abstract

**Background:** The COVID-19 pandemic led to social and physical distancing measures that reduced social contact and support. We explored whether people with more frequent and supportive social contact had fewer depressive symptoms during the UK Spring 2020 ‘lockdown’, whether this applied to face-to-face and remote electronic contact, and whether people with higher empathy levels, or more frequent pre-COVID social contact with others were more protected.

**Methods:** UK dwelling participants aged ≥18 in the internet-based longitudinal COVID-19 Social Study completed up to 22 weekly questionnaires about frequency of face-to-face and phone/video social contact, perceived social support, and depressive symptoms assessed with the patient health questionnaire (PHQ-9). Mixed linear models examined associations between social contact and support, and depressive symptoms. We examined for interaction by empathic concern and perspective taking and pre-COVID social contact frequency.

**Results:** In 71,117 people with mean age 49 years (standard deviation 15) we found that daily face-to-face or phone/video contact was associated with lower PHQ-9 scores (mean difference 0.258 (95% confidence interval 0.225, 0.290) and 0.117 (0.080, 0.154) respectively) compared to having no contact. Those with high social support scored 1.836 (1.801, 1.871) PHQ-9 points lower than those with low support. The odds ratio for depression for those with daily face-to-face social contact compared to no face-to-face contact was 0.712 (0.678, 0.747). Daily compared to no phone/video contact was associated with odds ratio for depression 0.825 (0.779, 0.873). And reporting high, compared to low, social support was associated with 0.145 (95%CI 0.138, 0.152) odds ratio for depression. The negative association between social relationships and depressive symptoms was stronger for those with high empathic concern, perspective taking and usual sociability.

**Conclusions:** Those who had more face-to-face contact during lockdown had fewer depressive symptoms. Phone or video communication were beneficial but less so. People who were usually more sociable or had higher empathy were more likely to have depressive symptoms during enforced reduced social contact. Results have implications both for our management of COVID-19 and potential future pandemics, and for our understanding of the relationship between social factors and mental health.

## Background

The novel coronavirus (2019-nCoV) related disease (COVID-19) [1] spread globally during early 2020, leading the World Health Organisation to declare an international public health emergency on 30^th^ January 2020 and a pandemic on 11^th^ March 2020. Its highly infectious nature and the effect on individuals and health services caused many countries to initiate physical and social distancing measures and close non-essential businesses and services. On 16^th^ March 2020, people in the UK were advised against all unnecessary social contact which included avoiding pubs, clubs, cinemas and restaurants [2]. One week later on 23^rd^ March 2020 a ‘lockdown’ was announced in the UK [3]. These regulations specified that people should not leave home except for once-daily exercise, medical needs, essential shopping or work. Gatherings of more than two people were prohibited. This legislation was changed on 1^st^ June 2020 when people living alone were permitted to meet with one other person outside, again on 13^th^ June when two single-adult households were allowed to pair with one another and have unlimited contact, and on 4^th^ July two households could to meet indoors and multiple households could meet outdoors.

The potential detrimental impact on mental health of changes in social relationships resulting from the lockdown, including reduced frequency of social contact and insufficient social support, has been acknowledged [4] and noted in previous pandemic quarantines [5]. Social relationships can be measured in several ways, and commonly is divided into structural social relationships (i.e. the number and type of people with whom a person interacts), and functional aspects (meaning the qualitative experience of those interactions, such as perception of social support or loneliness) [6]. Having better structural or functional social relationships is linked to improved mental health. A systematic review of observational studies found that perceived support from others was associated with lowered risk of depression and depressive symptoms [7]. Another added that having more extensive social networks was associated with lower risk of depression [8]. These associations are found throughout the life-course including in older people [8] and children and adolescents [9].

The experience of social relationships changed during the COVID-19 pandemic. People turned to remote methods of communication, with increased use of phone calls - for example average call duration increased from 3.7 to 5.4 minutes from January to April 2020 in the UK [10] - and higher use of mobile messaging and video-calling in the UK and throughout Europe[11]. It is important to consider whether these approaches to maintain relationships were associated with better mental health, as this may guide policy to reduce the impact of future periods of lockdown during COVID-19 and other pandemics.

Further, whilst there is evidence that richer social relationships are beneficial for mental health, it is unclear whether the unique circumstances of the COVID-19 pandemic, where reduction in social contact was imposed on all UK citizens, impacted the effect of social isolation on mental health. Socially active people may have suffered more from having social contact involuntarily reduced than those who had infrequent social contact prior to the pandemic. Furthermore, as reduced social contact was imposed on all for the purpose of promoting public health, it may have meant that psychological ability to think of others and conceive of the ‘greater good’ affected the experience of the lockdown. Higher empathic concern for others is associated with worse mental health, while higher ability to take the perspective of others is associated with better mental health [12, 13], so empathy may have moderated the effect of impaired social relationships on mental health, potentially creating high risk groups for psychiatric distress during periods of lockdown.

Therefore, we aimed to test, in a large prospective study initiated at the start of the UK lockdown, our hypotheses that poor structural social relationships measured by frequency of face-to-face and phone or video contact and functional social relationships (measured by perceived social support) would be associated with more depressive symptoms and higher risk of depression. We used data with repeated weekly measures of social relationships (exposure) and depressive symptoms (outcome) to give more robust estimates of the associations and capture changes in social relationships as lockdown regulations changed. Additionally, we explored whether these associations would be strengthened in those with previous high sociability and high levels of empathic concern, and ameliorated in those with higher perspective-taking ability.

## Methods

### Study design and participants

UCL Research Ethics Committee [12467/005] approved the study and all participants gave informed consent. We conducted a longitudinal analysis of data from the COVID-19 Social Study [14] of UK-dwelling participants aged 18 years and older. The COVID-19 Social Study started on 21^st^ March 2020 to consider social and mental effects of the COVID-19 pandemic in the UK at the beginning of the lockdown. The study was promoted through several routes: large databases of people who had consented to be contacted about health research; United Kingdom Research and Innovation mental health research networks; media coverage; and targeted recruitment to increase representativeness to people from low income, low educational, unemployed backgrounds, and vulnerable groups. Full details of the study protocol are available at www.covidsocialstudy.org.

Participants were invited by email to complete online questionnaires using the Redcap online survey tool (https://www.project-redcap.org/). They could enrol in the study at any time after 21^st^ March 2020 and, following their first extensive baseline questionnaire, received a weekly email until 21^st^ August with a link to a shorter follow-up questionnaire. Participants who did not complete the weekly questionnaire received two reminder emails. If they still did not respond to the survey then they were counted as lost to follow-up.

Eligibility criteria for participants in our study were 1) being aged ≥18 years, 2) joining the COVID-19 study any time between study inception on 21^st^ March 2020 and 21^st^ August 2020, the last date of weekly data collection, and 3) residing in the UK at the time of baseline questionnaire completion.

### Measures

#### Social relationship variables

Data about social relationships were collected weekly. For structural relationships, we asked:

1. The number of days during the past week on which participants had at least 15 minutes of face-to-face social contact (including with those with whom they live).
2. The number of days during the past week on which participants had at least 15 minutes of telephone or video social contact.

We treated face-to-face and virtual contact separately, as responses to the two questions did not correlate (*r* = 0.05). Responses were 0-7, and we considered this as a continuous scale, and also categorised into none (0 days), some (1-6 days) and daily (7 days).

For functional relationships we used an adapted version of the six item short form of Perceived Social Support Questionnaire, which was administered weekly. It has good psychometric properties [15] and we made minor adaptations to make the language more relevant to experiences during COVID-10 pandemic (appendix 1). Participants were asked to rate their agreement on a 5 point Likert scale from ‘not at all true’ to ‘very true’ with six statements about their feelings during the past week. Higher scores indicated more perceived social support. We considered this as a continuous variable and categorised scores into tertiles based on the distribution of the group (low <3, medium = 3 to 3.99, high ≥4.)

### Depressive symptoms

We measured depressive symptoms weekly using the Patient Health Questionnaire (PHQ-9) [16], a standard instrument for assessing the severity of depressive symptoms in primary care settings. The questionnaire involves nine items, with responses on a Likert scale of 0 to 3 ranging from ‘not at all’ to ‘nearly every day’ with higher scores indicating more depressive symptoms. We used the scale as continuous, and also generated a binary variable as depressed / not depressed using a cut-off point of ≥10 [17].

### Potential moderators

#### Empathic concern and perspective taking

We used 14 questions from the Interpersonal Reactivity Index (IRI) [18] to evaluate self-reported empathic concern and perspective taking in one of the weekly surveys during week 13 of the study (13^th^-20^th^ June 2020). Empathic concern and perspective taking have low-moderate correlation [18] and differential associations with depressive symptoms [12] so were treated separately in our analyses. The IRI is validated in the general population and is a measure of ‘trait-based’ empathy aiming to assess long-term, rather than situational, empathic responses, so does not focus on feelings related to the COVID-19 pandemic [19]. Participants were asked to rate their agreement with statements on a 5-point Likert scale ranging from “Does not describe me well” to “Describes me very well”. Scores for the two subscales were averaged across each domain giving mean scores for empathic concern and perspective taking. We used this mean score and categorised tertiles based on the distribution in the sample (low <3.7, medium 3.7 to 4.19, high ≥4.2 for empathic concern, and low <3.3, medium 3.3 to 3.99, high ≥4 for perspective taking).

#### Usual social contact

We measured social contact prior to the COVID-19 pandemic at baseline with the question ‘Usually in your life, how often do you meet up with people face to face socially, not for work (e.g. friends, family, relatives or social events with colleagues)?’ Participants chose from response options ‘less than once weekly’, ‘once or twice per week’, or ‘three or more per week’.

### Confounders

We used other variables from the baseline interview which we considered from previous evidence to be potential confounders: age; gender (male, female, other/prefer not to say); ethnicity (White, other); highest educational attainment (lower secondary (GCSE/O-level or lower), higher secondary (A-level or equivalent), graduate or higher); living alone or living with others; marital status (cohabiting with partner or spouse, partner or spouse but living apart, divorced or widowed, single and never married); in employment/study or retired/not working; annual household income less or more than £30,000.

### Analysis

We summarised the demographic characteristics of our sample and on the social relationship variables, and their correlation using Spearman’s rank test, and depression scale, as well as the potential moderators of empathic concern, perspective taking and usual pre-COVID social contact.

#### Association of social relationships with depressive symptoms and depression

We used mixed linear models with a random effect for intercept because data were clustered by individuals within study week. This method is preferable to statistical approaches that take just a single cross-sectional time-point for each individual as they allowed us to use all available exposure (social relationships) and outcome (PHQ-9) weekly data within the period of data collection which was appropriate to allow our analysis to account for the variation in lockdown restrictions during the study.

We constructed models examining the association, in separate models, of face to face contact, phone/video contact, or social support with depressive symptoms, which were initially unadjusted (model 1), adjusted for age and sex (model 2), and additionally adjusted for education, employment status and income (model 3). A final model (model 4) additionally adjusted for amount of social contact (time-varying), meaning that the model examining face-to-face contact additionally adjusted for phone/video contact, the model with phone/video contact additionally adjusted for face-to-face contact, and the model for social support adjusted for both face-to-face and phone/video contact. We examined the social relationship variables as both continuous and categorical. We repeated these analyses in mixed effect logistic regression models with depression as a binary outcome with a cut-point from PHQ-9 of 10, adjusted as in the linear regression models.

#### Moderation by empathy and usual social contact

We repeated model 3 of the analysis of social relationships (treated as continuous variables) and depressive symptoms with the potential moderating variables of empathic concern and perspective taking (as continuous variables) and usual social contact (as a categorical variable) and interaction terms of these with the social relationship variables (continuous) in separate models with depressive symptoms outcomes. Where there was statistical evidence of interaction (p<0.05), we presented results for the association between the social relationship variables and depressive symptoms stratified according to tertiles of empathic concern, perspective taking, and the three categories of usual social contact.

## Results

We included 71,117 participants who answered 679,615 weekly questionnaires between 21^st^ March and 21^st^ August 2020; Table 1 details their demographic characteristics. Three quarters (53,026) were women and the mean age was 49 years (standard deviation (SD) 15). 66,673 (93.8%) were from White ethnic groups and 47,652 (67.0%) had attained graduate or higher educational level. 44,863 (63.1%) were married or cohabiting with their partner or spouse and 12,735 (17.9%) were living alone. 46,333 (65.2%) were in employment or full-time education with the remainder not employed or retired.

**Table 1:** Characteristics of the sample (N=71, 117)

| Characteristic | Category | n/N (%) or* Mean [41] |
| --- | --- | --- |
| **Age (years)** | Mean [41] | 49.1 (14.9) |
|  | 18-24 | 2,872 (4.0) |
|  | 25-34 | 10,858 (15.3) |
|  | 35-44 | 14,521 (20.4) |
|  | 45-54 | 15,602 (21.9) |
|  | 55-64 | 14,426 (20.3) |
|  | 65-74 | 10,230 (14.4) |
|  | ≥75 | 2,608 (3.7) |
| **Gender** | Female | 53,026 (74.6) |
|  | Male | 17,756 (25.0) |
|  | Other / prefer not to say | 335 (0.5) |
| **Ethnicity** | White | 66,673 (93.8) |
|  | Other | 4,179 (5.9) |
|  | *Missing* | 265 (0.4) |
| **Educational level** | Lower secondary | 10,576 (14.9) |
|  | Higher secondary | 12,889 (18.1) |
|  | Graduate | 47,652 (67.0) |
| **Living situation** | Lives alone | 12,735 (17.9) |
|  | Lives with others | 58,382 (82.1) |
| **Marital status** | Cohabiting with partner/spouse | 44,863 (63.1) |
|  | Living apart from partner/spouse | 4,903 (6.9) |
|  | Divorced/widowed | 8,973 (12.6) |
|  | Single, never married | 12,378 (17.4) |
| **Employment** | In employment | 46,333 (65.2) |
|  | Retired / not working | 24,784 (34.8) |
| **Household income** | < £30,000 | 25,141 (35.4) |
|  | ≥ £30,000 | 39,422 (55.4) |
|  | *Prefer not to say* | 6,554 (9.2) |
| **Usual social contact** | Less than once weekly | 21,171 (29.8) |
|  | Once or twice per week | 24,239 (34.1) |
|  | Three or more per week | 25,428 (35.8) |
|  | *Missing* | 279 (0.4) |
| **Mean interpersonal reactivity index scores** | Empathic concern (n=29,437) | 4.0 (0.7) |
|  | Perspective taking (n=29,573) | 3.7 (0.7) |

**Note:** SD = standard deviation.

Of the 69,975 participants (Table 2) who gave baseline information about face-to-face contact, the median number of days with face-to-face contact at baseline was 7 (interquartile range (IQR) 3, 7), with 44,676 (63.9%) having daily contact and 10,476 (15.0%) having no face-to-face contact during the preceding week. The median number of days on which participants (n=70,074) had 15 minutes or more of phone/video contact was 4 (IQR 2,7) with 5,964 (8.5%) reporting no days, 43,022 (61.4%) 1-6 days and 21,088 (30.1%) reporting daily phone/video contact. Median social support score was 4.0 (IQR 3.2, 4.5) in the 68,784 who completed this questionnaire at baseline. The correlation between face-to-face and phone/video contact was 0.05; between face-to-face and social support was 0.27; and between phone/video contact and social support was 0.21. Mean PHQ-9 score was 7.1 (SD 6.1) and 20,235 of 69,671 participants (29.0%) scored 10 or more.

**Table 2:**
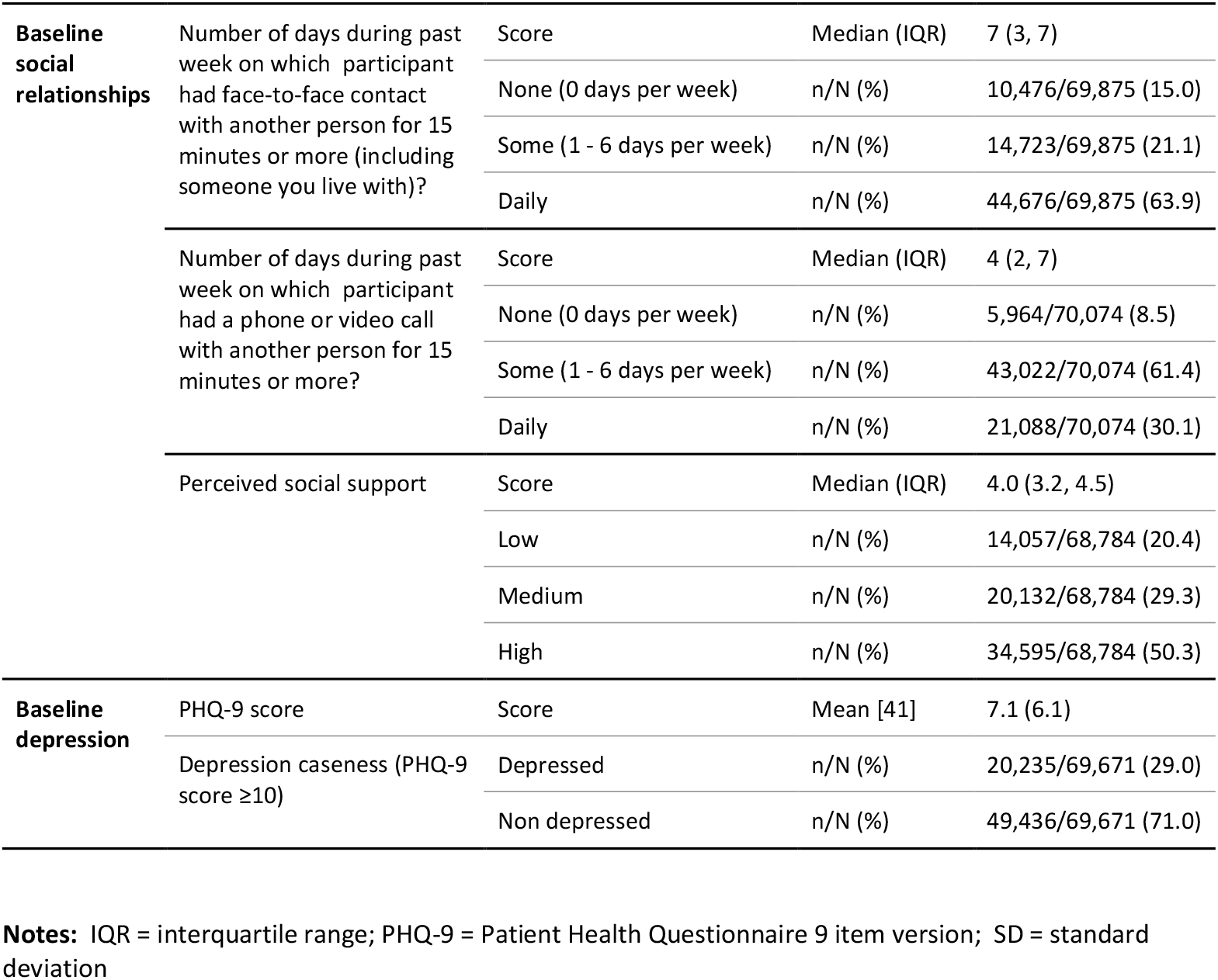
Baseline structural and functional social relationships and depression

### Association of social relationships with depressive symptoms and depression

Results from our analyses of the association between social relationships and depressive symptoms are summarised in Table 3. Each additional day of face-to-face contact was associated with 0.052 (95% confidence interval (CI) 0.048, 0.057) lower depression score adjusted for socio-demographic characteristics, and 0.051 (95%CI 0.047, 0.056) when additionally adjusted for phone/ video contact. Having daily contact was associated with 0.258 (95%CI 0.225, 0.290) lower PHQ-9 score than having no face-to-face contact, (0.247 (0.215, 0.280) when adjusted for phone/video contact).

**Table 3.** Association of structural and functional social relationships with depressive symptoms

|  |  | **Model 1** |  | **Model 2** |  | **Model 3** |  | **Model 4** |  |
| --- | --- | --- | --- | --- | --- | --- | --- | --- | --- |
|  |  | **unadjusted** |  | **Adjusted for age and sex** |  | **Model 2 + adjusted for education, employment status and income** |  | **Model 3 + adjusted for face-to-face and/or phone/video contact** |  |
|  |  | Coefficient | (95% CI) | Coefficient | (95% CI) | Coefficient | (95% CI) | Coefficient | (95% CI) |
| **Number of days during past week on which participant had face-to-face contact with another person for 15 minutes or more** | Per 1 day more contact | **-0.051** | **(-0.056, -0.047)** | **-0.059** | **(-0.064, -0.055)** | **-0.052** | **(-0.057, -0.048)** | **-0.051** | **(-0.056, -0.047)** |
|  | None (0 days per week) | reference |  | reference |  | reference |  | reference |  |
|  | Some (1 - 6 days per week) | -0.003 | (-0.031, 0.026) | -0.021 | (-0.049, 0.008) | -0.012 | (-0.042, 0.018) | -0.010 | (-0.040, 0.020) |
|  | Daily | **-0.251** | **(-0.282, -0.220)** | **-0.302** | **(-0.333, -0.271)** | **-0.258** | **(-0.290, -0.225)** | **-0.247** | **(-0.280, -0.215)** |
| **Number of days during past week on which participant had phone or video call contact with another person for 15 minutes or more** | Per 1 day more contact | **-0.030** | **(-0.034, -0.025)** | **-0.033** | **(-0.038, -0.029)** | **-0.026** | **(-0.031, -0.021)** | **-0.023** | **(-0.028, -0.018)** |
|  | None (0 days per week) | reference |  | reference |  | reference |  | reference |  |
|  | Some (1 - 6 days per week) | **-0.078** | **(-0.106, -0.049)** | **-0.094** | **(-0.122, -0.066)** | **-0.061** | **(-0.090, -0.031)** | **-0.043** | **(-0.073, -0.013)** |
|  | Daily | **-0.162** | **(-0.196, -0.127)** | **-0.186** | **(-0.220, -0.151)** | **-0.145** | **(-0.182, -0.108)** | **-0.117** | **(-0.154, -0.080)** |
| **Perceived social support** | Per one point increase | **-1.020** | **(-1.034, -1.005)** | **-1.057** | **(-1.071, -1.043)** | **-1.033** | **(-1.048, -1.018)** | **-1.035** | **(-1.050, -1.020)** |
|  | Low | reference |  | reference |  | reference |  | reference |  |
|  | Medium | **-0.939** | **(-0.967, -0.912)** | **-0.983** | **(-1.010, -0.955)** | **-0.962** | **(-0.991, -0.933)** | **-0.956** | **(-0.986, -0.927)** |
|  | High | **-1.819** | **(-1.851, -1.786)** | **-1.890** | **(-1.922, -1.857)** | **-1.848** | **(-1.882, -1.814)** | **-1.836** | **(-1.871, -1.801)** |

**Notes:** Coefficient represents mean number of points on PHQ-9 higher or lower according to social relationship characteristic

Each additional day of phone/video contact was associated with 0.026 (95%CI 0.021, 0.031) lower PHQ-9 score (0.023 (95%CI 0.018, 0.028) when adjusted for amount of face-to-face contact). Having some or daily phone or video contact was associated with lower depressive symptoms compared to no phone or video contact when adjusted for face-to-face contact; PHQ-9 scores 0.043 (95%CI 0.013, 0.073), and 0.117 (95%CI 0.080, 0.154) lower for some and daily contact respectively compared to no phone/video contact.

Higher reported perceived social support was associated with lower depressive symptoms: for each additional point, PHQ-9 score was 1.033 (95%CI 1.018, 1.048) lower in models adjusting for sociodemographic characteristics and 1.035 (95%CI 1.020, 1.050) when additionally adjusting for amount of face-to-face and phone/video contact. Those in the highest social support tertile scored a mean 1.836 (95%CI 1.801, 1.871) points lower on PHQ-9 than those in the lowest tertile in fully adjusted models.

A similar pattern of results was seen in multilevel logistic regression models with a binary depression outcome (Table 4). The odds ratio for depression for people having daily compared to no face-to-face social contact was 0.712 (95%CI 0.747, 0.678) in models adjusting for sociodemographic characteristics and phone/video contact. Those with daily phone/video contact compared to no contact had odds ratio for depression 0.825 (95%CI 0.779, 0.873) in fully adjusted models. Reporting high, compared to low, social support was associated with 0.145 (95%CI 0.138, 0.152) odds ratio for depression, adjusted for sociodemographic status and face-to-face and phone/video contact.

**Table 4.** Association of structural and functional social relationships with depression (Odds ratio for depression)

|  |  | Model 1<br>unadjusted |  | Model 2<br>Adjusted for age and sex |  | Model 3<br>Model 2 + adjusted for<br>education, employment<br>status and income |  | Model 4<br>Model 3 + adjusted for<br>face-to-face and/or<br>phone/video contact |  |
| --- | --- | --- | --- | --- | --- | --- | --- | --- | --- |
|  |  | Odds Ratio | (95% CI) | OR | 95% CI | OR | 95% CI | OR | 95% CI |
| **Number of days during past week on which participant had face-to-face contact with another person for 15 minutes or more** | Per 1 day more contact | **0.937** | **(0.931, 0.943)** | **0.921** | **(0.915, 0.927)** | **0.937** | **(0.931, 0.944)** | **0.939** | **(0.933, 0.945)** |
|  | None (0 days per week) |  | reference |  | reference |  | reference |  | reference |
|  | Some (1 - 6 days per week) | **0.954** | **(0.913, 0.997)** | **0.912** | **(0.873, 0.954)** | **0.945** | **(0.903, 0.990)** | **0.945** | **(0.903, 0.990)** |
|  | Daily | **0.701** | **(0.669, 0.734)** | **0.625** | **(0.596, 0.655)** | **0.703** | **(0.669, 0.738)** | **0.712** | **(0.678, 0.747)** |
| **Number of days during past week on which participant had phone or video call contact with another person for 15 minutes or more** | Per 1 day more contact | **0.949** | **(0.942, 0.955)** | **0.942** | **(0.935, 0.948)** | **0.956** | **(0.949, 0.963)** | **0.959** | **(0.952, 0.966)** |
|  | None (0 days per week) |  | reference |  | reference |  | reference |  | reference |
|  | Some (1 - 6 days per week) | **0.861** | **(0.823, 0.900)** | **0.830** | **(0.794, 0.868)** | **0.890** | **(0.849, 0.932)** | **0.906** | **(0.864, 0.949)** |
|  | Daily | **0.777** | **(0.736, 0.819)** | **0.737** | **(0.700, 0.778)** | **0.799** | **(0.755, 0.845)** | **0.825** | **(0.779, 0.873)** |
| **Perceived social support** | Per one point increase | **0.349** | **(0.342, 0.355)** | **0.332** | **(0.326, 0.338)** | **0.356** | **(0.349, 0.363)** | **0.357** | **(0.350, 0.364)** |
|  | Low |  | reference |  | reference |  | reference |  | reference |
|  | Medium | **0.387** | **(0.372, 0.402)** | **0.362** | **(0.349, 0.376)** | **0.387** | **(0.372, 0.403)** | **0.393** | **(0.377, 0.409)** |
|  | High | **0.137** | **(0.131, 0.144)** | **0.122** | **(0.117, 0.128)** | **0.141** | **(0.134, 0.147)** | **0.145** | **(0.138, 0.152)** |

### Moderation by empathy and usual social contact

We repeated analyses with interaction terms for potential moderators, results are described in Figure 1, which shows the number of points difference on PHQ-9 depression scale associated with one point higher social relationship score, stratified by level of empathic concern, perspective taking and social contact. We found an interaction for both empathic concern and perspective taking in the negative relationship between face-to-face contact and depressive symptoms, with a higher association in participants with higher empathy. In the 29,567 people who completed the empathic concern section of the IRI, those with high empathic concern scored 0.068 (95%CI 0.058, 0.077) points lower on PHQ-9 for each additional day of social contact, compared to those with low empathic concern who scored 0.041 (95%CI 0.033, 0.050) lower with more social contact. In the 29,701 people who completed the perspective taking questionnaire, participants with high perspective taking scores scored 0.075 (95%CI 0.066, 0.084) points lower on depressive symptoms for each additional day of face-to-face contact, whereas those with low perspective taking scored 0.038 (95%CI 0.029, 0.047) points lower with more face-to-face contact. Higher empathic concern and perspective taking however was linked to smaller association between perceived social support and depressive symptoms. Those with higher empathic concern and perspective taking reported lower depressive symptoms with additional social support, compared to those with lower scores on the empathy variables.

**Figure 1.**
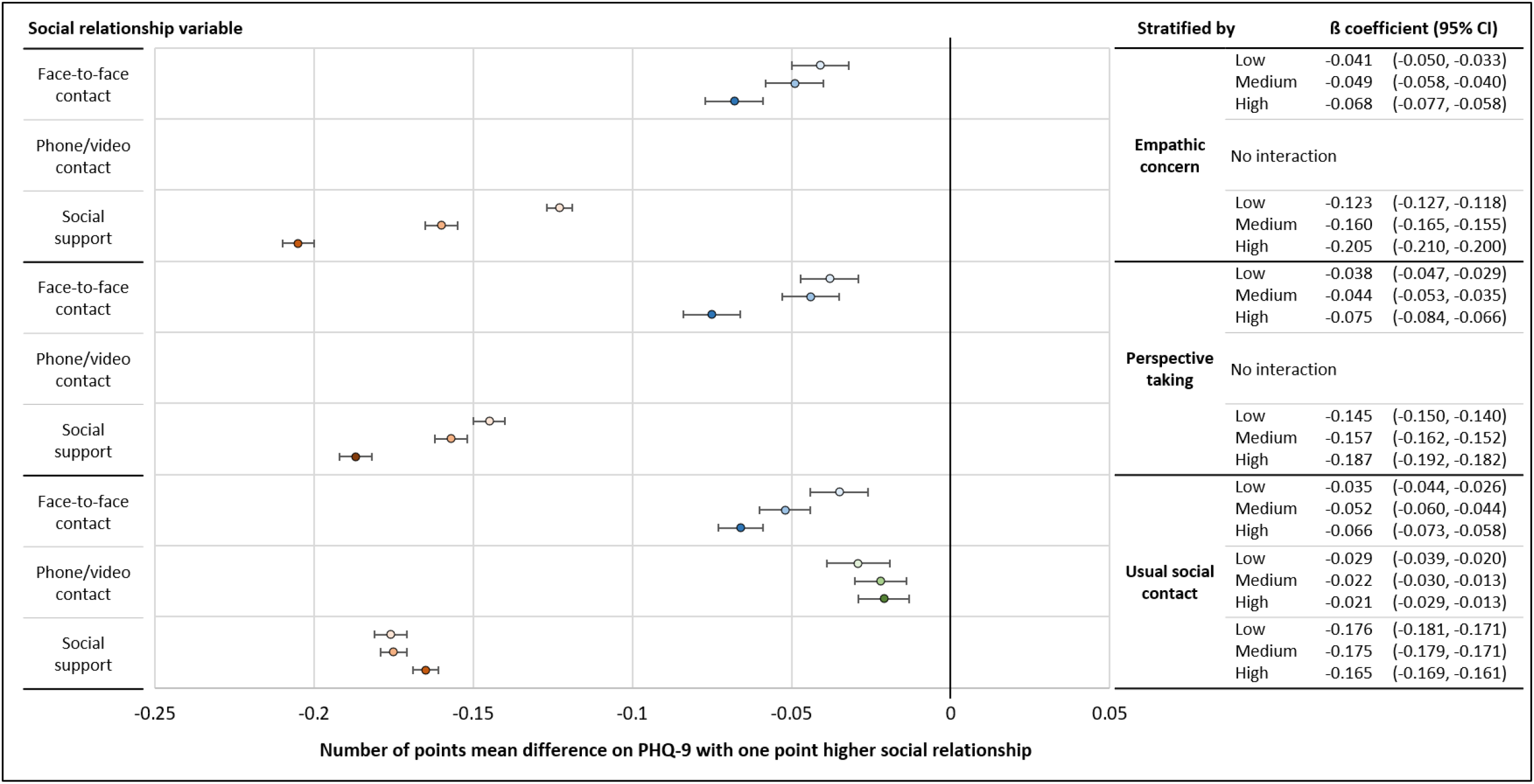
Association of face-to-face contact, phone/video contact and social support with depressive symptoms, stratified by empathic concern, perspective taking and usual social contact **Notes:** Coefficients indicate the number of points difference on PHQ-9 depression scale associated with one point higher social relationship score, stratified by level of empathic concern, perspective taking and social contact. All analyses are adjusted for age, sex, education, employment status and income.

We found evidence of interaction between pre-COVID social contact and the relationship between face-to-face contact and depressive symptoms. Those who usually had three or more social meetings per week scored 0.066 (95%CI 0.058, 0.073) points lower on the depression scale with each extra day of social contact during the lockdown, whereas those who usually met socially with people less than once weekly experienced less effect on depression with social contact, scoring 0.035 (95%CI 0.026, 0.044) points lower on the depression scale for each additional social contact. Small differences were seen in the associations between phone/video contact and depressive symptoms and social support and depressive symptoms according to level of usual pre-COVID social contact. In these analyses those who usually had more social contact experienced a marginally smaller effect of having more phone/video contact or social support on depressive symptoms than those who usually had infrequent social contact.

## Discussion

This study explored how structural and functional aspects of social relationships, measured by the frequency of face-to-face and phone/video contact and level of perceived social support from others, were associated with depressive symptoms during the COVID-19 related lockdown in the UK. We found that both more frequent face-to-face and phone/video contact were associated with lower depressive symptoms, including when mutually adjusted for one another, and in-person contact had more effect than digital contact. Those with higher levels of perceived social support had lower depressive symptoms and depression risk. These relationships persisted when adjusted for the amount of actual weekly face-to-face or digital social contact.

Our main finding that having more or better social relationships was associated with lower depressive symptoms and risk of depression is consistent with the literature. A systematic review of 51 studies, of which 23 were prospective, reported strong consistent findings that having more perceived support from others and more extensive social networks was associated with lower risk of depression [8]. Establishing the direction of this association can be challenging. Many studies suggest that having positive and enjoyable social experiences and recalling socially rewarding information is linked to lower depressive symptoms [20], that experiencing loneliness is linked to higher threat vigilance which may promote depressive cognitions [21], and that social support may moderate the effect of stressful life events on health [22]. This has been confirmed through studies tracking experiences longitudinally, including a recent study of UK older adults that found greater risk of depression up to 12 years later in those reporting loneliness [23]. However, the association may be bidirectional as depressive symptoms such as reduced capacity for enjoyment, interest and concentration, and impaired self-esteem and self-confidence may impair social relationships or a person’s perception of them. Therefore, associations between self-rated structural and functional aspects of relationships may partly be a manifestation of depressive symptomatology. What is unique in this study is that quantity of social relationship was affected by law for everyone in the UK. While people who are depressed may usually decide not to see people socially, during this period everyone had their social contact restricted.

Our findings also have specific relevance to the social context of the COVID-19 pandemic, echoing some previous studies. For example, one cross-sectional study of 7,127 UK older adults aged 70 on average who self-reported whether their mental health had changed, found that loneliness was associated with reporting a worsening of depression and anxiety symptoms, although this study did not objectively identify change in mental health [24]. In our study, the association was relatively modest; the minimum difference on the PHQ-9 considered to indicate a clinically meaningful improvement in response to treatment being between 2.8 and 5 points [25]. Associations with depression were stronger for the functional rather than structural measure of social relationships and persisted when adjustment for structural relationships was included. This may indicate that it is the quality, rather than quantity, of social relationships which matters most, or may reflect the overlap between depressive symptomatology and negative judgment about support from others. Importantly, our study also provided new findings on the relationship between face-to-face vs virtual communication and mental health.

Some studies conducted prior to the COVID-19 pandemic have suggested similar benefits of video contact. In a study of young adults, both face to face and video enjoyable interactions were associated with improvements in self-esteem but face-to-face interaction had a stronger effect [26]. A further study of US college students reported that affect improved more following in-person communication than digital communication [27]and another study reported that relationship strength was higher following face-to-face communication compared to video, audio and text communication [28]. Our study suggests that these experimental findings apply in a pandemic-enforced lockdown. Face-to-face contact appeared to confer greater benefit, but when in-person communication was taken into account, remote communication remained beneficial. Use of social media for communication is common, with 88% of young adults reporting using it in 2018 [29], and use has increased during the COVID-19 pandemic [10]. Social information processing theory suggests that frequent use of digital communication over time allows users to convey and process personal information effectively, despite fewer nonverbal cues than in face-to-face settings [30] so remote communication is likely to be beneficial. It may also be that our study’s measure of face-to-face social contact during the lockdown captured contact with close family and friends, as contact was most likely with cohabiting people, whereas digital contact was more likely to be with more distant contacts. Therefore the stronger association of in-person rather than digital communication may have partly reflected the relationship, rather than the method of contact.

A secondary aim of our study was to explore whether having more empathy for others or usual sociability affected these associations; the first known study to do this. We found that higher empathic concern for others was linked to a stronger effect of structural and functional social relationships on increasing depression, which is consistent with the associations between higher empathic concern and depressive symptoms [31]. Deprivation of social relationships may be felt more strongly for those who empathise more with others. Contrary to our hypothesis, higher perspective taking did not attenuate the detrimental effect of social isolation on mental health and instead, strengthened the relationship. Our hypothesis was based on studies reporting an inverse relationship between perspective taking and depressive symptoms, usually in caring groups such as family carers and healthcare workers [12, 13, 32, 33], suggesting that ability to conceptualise another’s perspective may moderate the experience of their distress. However, our finding may indicate that people who had others’ perspectives in mind found the lockdown more difficult as, in these circumstances, they had little agency to act to alleviate others’ situations.

Finally, we report a novel finding that the effect of impaired structural and functional relationships during the lockdown was associated more strongly with depression in people who were previously more socially active. Several studies have suggested that unmarried people who live alone have wider and more active social networks and these have a greater impact on wellbeing for single people than for those in relationships [34-36]. In our sample, those with less daily face-to-face contact were likely to be living alone and therefore unable to mix socially with the wide range of people with whom they would normally do so, and this may have been detrimental to mental health.

### Limitations

While our large sample size covered an extensive range of sociodemographic characteristics, it was not nationally representative with some groups being underrepresented, for example those from lower sociodemographic groups and minority ethnic groups. However, the potential bias in selection is less relevant for examining risk factor-outcome associations [37]. All variables were by self-report, so negative perspectives common in depression may have influenced report of structural and functional relationships, which would likely overestimate the association.

The questionnaire design, whereby respondents had to answer all questions to proceed to the next page, meant that there was little missing data for the different questionnaire domains, but participation varied longitudinally, with around 10% answering only one questionnaire, and only 10% answering all the weekly questionnaires. Our analysis did not account for attrition which may have been higher in those with depressive symptoms, and we could only examine moderation by empathy in the smaller sample of participants who answered that weekly questionnaire. Participants also joined at different stages, with around 40% joining within the first week of the survey in late March 2020, and others joining at any subsequent point. Our analytic approach allowed us to make use of all repeated exposure and outcome variables, and this was particularly relevant for the circumstances of lockdown whereby social contact with others could vary markedly from week-to-week as new legislation came into force, but our approach makes it difficult to be certain of the direction of association and reverse causality may have affected our results. Finally, we lacked detail about the nature of social interactions, such as the duration and intensity of social contact and there is the potential for residual confounding from unmeasured confounders.

### Clinical and research implications

This large longitudinal study of structural and functional relationships and depressive symptoms throughout the first COVID-19 related lockdown in the UK supports the existing literature that both structural and functional aspects of social relationships are associated with better mental health. Our study adds that good quality and supportive face-to-face contact with others is likely to be most beneficial but that, even when this is not available or permitted, phone and video contact may be beneficial. We also find that the impact of social isolation may be most hard-felt for those who are usually socially active and more empathic.

These findings have immediate clinical and public health relevance. The UK has already had further periods of physical and social distancing due to COVID-19 and these are likely in the future, so identifying high risk groups for negative effects is important. Social isolation is associated with other adverse cognitive [38] and physical effects [39], so public health policy should facilitate social contact, where possible, to alleviate the burden on mental health especially for those who live alone and are accustomed to contact with others. Individuals should use digital methods of communication when in-person meetings are limited. There is need for actions to improve social connectedness throughout this and potential future pandemics [40] to reduce the potential for mental illness arising from social isolation.

## Supporting information

Strobe Checklist

## Data Availability

Data is from the COVID-19 Social Study which is currently not publicly available, but interested collaborators may contact the study team.

## Acknowledgements

The researchers are grateful for the support of several organisations with their recruitment efforts including: the UKRI Mental Health Networks, Find Out Now, UCL BioResource, SEO Works, FieldworkHub, and Optimal Workshop. The study was also supported by HealthWise Wales, the Health and Care Research Wales initiative, which is led by Cardiff University in collaboration with SAIL, Swansea University.

## Ethical standards

The authors assert that all procedures contributing to this work comply with the ethical standards of the relevant national and institutional committees on human experimentation and with the Helsinki Declaration of 1975, as revised in 2008. The study was approved by the UCL Research Ethics Committee [12467/005] and all participants gave informed consent.

## Appendix 1 Comparison of items in the original and revised Perceived Social Support Questionnaire

Appendix 1 – Comparison of items in the original and revised Perceived Social Support Questionnaire
| Original | Adapted for COVID-19<br>In the past week, I feel... |
| --- | --- |
| I experience a lot of understanding and security from others | I have experienced a lot of understanding and support from others |
| I know a very close person whose help I can always count on | I have a very close person whose help I can always count on |
| If necessary, I can easily borrow something I might need from neighbours or friends | If necessary, I can easily borrow something I need from neighbours or friends |
| I know several people with whom I like to do things | I have people with whom I can spend time and do things together |
| When I am sick, I can without hesitation ask friends and family to take care of important matters for me | If I get sick, I have friends and family who will take care of me |
| If I am down, I know to whom I can go without hesitation | If I am feeling down, I have people I can talk to without hesitation |

## References

1. Zhu N, Zhang D, Wang W, Li X, Yang B, Song J, et al. A novel coronavirus from patients with pneumonia in China, 2019. New England Journal of Medicine. 2020.

2. The Health Foundation. COVID-19 policy tracker 2020 [cited 2020 06/11/2020]. Available from: https://www.health.org.uk/news-and-comment/charts-and-infographics/covid-19-policy-tracker.

3. UK Government Cabinet Office. Guidance: Staying at home and away from others (social distancing) 2020 [cited 2020 06/11/2020]. Available from: https://www.gov.uk/government/publications/full-guidance-on-staying-at-home-and-away-from-others/full-guidance-on-staying-at-home-and-away-from-others.

4. Luykx JJ, Vinkers CH, Tijdink JK. Psychiatry in Times of the Coronavirus Disease 2019 (COVID-19) Pandemic: An Imperative for Psychiatrists to Act Now. JAMA Psychiatry. 2020.

5. Brooks SK, Webster RK, Smith LE, Woodland L, Wessely S, Greenberg N, et al. The psychological impact of quarantine and how to reduce it: rapid review of the evidence. The Lancet. 2020.

6. Valtorta NK, Kanaan M, Gilbody S, Hanratty B. Loneliness, social isolation and social relationships: what are we measuring? A novel framework for classifying and comparing tools. BMJ open. 2016;6(4).

7. Gariepy G, Honkaniemi H, Quesnel-Vallee A. Social support and protection from depression: systematic review of current findings in Western countries. The British Journal of Psychiatry. 2016;209(4):284–93.

8. Santini ZI, Koyanagi A, Tyrovolas S, Mason C, Haro JM. The association between social relationships and depression: a systematic review. Journal of affective disorders. 2015;175:53–65.

9. Loades ME, Chatburn E, Higson-Sweeney N, Reynolds S, Shafran R, Brigden A, et al. Rapid Systematic Review: The Impact of Social Isolation and Loneliness on the Mental Health of Children and Adolescents in the Context of COVID-19. Journal of the American Academy of Child & Adolescent Psychiatry. 2020.

10. Ofcom. Mobile matters: Researching people’s experience of using Android mobile services. 2020 08/10/2020. Report No.

11. Sun S, Folarin AA, Ranjan Y, Rashid Z, Conde P, Stewart C, et al. Using Smartphones and Wearable Devices to Monitor Behavioral Changes During COVID-19. J Med Internet Res. 2020;22(9):e19992. Epub 25.9.2020. doi: 10.2196/19992. PubMed PMID: 32877352.

12. Tully EC, Ames AM, Garcia SE, Donohue MR. Quadratic associations between empathy and depression as moderated by emotion dysregulation. The Journal of Psychology. 2016;150(1):15–35.

13. Lee HS, Brennan PF, Daly BJ. Relationship of empathy to appraisal, depression, life satisfaction, and physical health in informal caregivers of older adults. Research in Nursing & Health. 2001;24(1):44–56. doi: 10.1002/1098-240x(200102)24:1<44::Aid-nur1006>3.0.Co;2-s.

14. Fancourt D, Steptoe A, Bu F. Trajectories of depression and anxiety during enforced isolation due to COVID-19: longitudinal analyses of 59,318 adults in the UK with and without diagnosed mental illness. medRxiv. 2020.

15. Kliem S, Mößle T, Rehbein F, Hellmann DF, Zenger M, Brähler E. A brief form of the Perceived Social Support Questionnaire (F-SozU) was developed, validated, and standardized. Journal of clinical epidemiology. 2015;68(5):551–62.

16. Kroenke K, Spitzer RL, Williams JB. The PHQ-9: validity of a brief depression severity measure. Journal of general internal medicine. 2001;16(9):606–13. Epub 2001/09/15. doi: 10.1046/j.1525-1497.2001.016009606.x. PubMed PMID: 11556941; PubMed Central PMCID: PMCPMC1495268.

17. Levis B, Benedetti A, Thombs BD. Accuracy of Patient Health Questionnaire-9 (PHQ-9) for screening to detect major depression: individual participant data meta-analysis. bmj. 2019;365.

18. Davis MH. A multidimensional approach to individual differences in empathy. 1980.

19. Konrath S. A critical analysis of the Interpersonal Reactivity Index. MedEdPORTAL Directory and Repository of Educational Assessment Measures (DREAM). 2013.

20. Lewis G, Kounali DZ, Button K, Duffy L, Wiles N, Munafo M, et al. Variation in the recall of socially rewarding information and depressive symptom severity: a prospective cohort study. Acta Psychiatrica Scandinavica. 2017;135(5):489–98.

21. Hawkley LC, Cacioppo JT. Loneliness matters: A theoretical and empirical review of consequences and mechanisms. Annals of behavioral medicine. 2010;40(2):218–27.

22. Cobb S. Social support as a moderator of life stress. Psychosomatic medicine. 1976.

23. Lee SL, Pearce E, Ajnakina O, Johnson S, Lewis G, Mann F, et al. The association between loneliness and depressive symptoms among adults aged 50 years and older: a 12-year population-based cohort study. The Lancet Psychiatry. 2020.

24. Robb CE, de Jager CA, Ahmadi-Abhari S, Giannakopoulou P, Udeh-Momoh C, McKeand J, et al. Associations of social isolation with anxiety and depression during the early COVID-19 pandemic: a survey of older adults in London, UK. Frontiers in Psychiatry. 2020;11.

25. Löwe B, Unützer J, Callahan CM, Perkins AJ, Kroenke K. Monitoring depression treatment outcomes with the patient health questionnaire-9. Medical care. 2004:1194–201.

26. Subrahmanyam K, Frison E, Michikyan M. The relation between face-to-face and digital interactions and self-esteem: A daily diary study. Human Behavior and Emerging Technologies. 2020;2(2):116–27.

27. Holtzman S, DeClerck D, Turcotte K, Lisi D, Woodworth M. Emotional support during times of stress: Can text messaging compete with in-person interactions? Computers in Human Behavior. 2017;71:130–9.

28. Sherman LE, Michikyan M, Greenfield PM. The effects of text, audio, video, and in-person communication on bonding between friends. Cyberpsychology: Journal of psychosocial research on cyberspace. 2013;7(2).

29. Pew Research Center. Social media use in 2018. 2018.

30. Walther JB. Theories of computer-mediated communication and interpersonal relations. In: Knapp M, Daly J, editors. The handbook of interpersonal communication. 4. Thousand Oaks, California, US: SAGE Publications; 2011. p. 443–79.

31. O’Connor LE, Berry JW, Lewis T, Mulherin K, Crisostomo PS. Empathy and depression: the moral system on overdrive. In: Farrow T, Woodruff P, editors. Empathy in mental illness. Cambridge, UK: Cambridge University Press; 007. p. 49–75.

32. Hollinger-Samson N, Pearson JL. The relationship between staff empathy and depressive symptoms in nursing home residents. Aging & Mental Health. 2000;4(1):56–65.

33. Jütten LH, Mark RE, Sitskoorn MM. Empathy in informal dementia caregivers and its relationship with depression, anxiety, and burden. International journal of clinical and health psychology. 2019;19(1):12–21.

34. Stokes JE, Moorman SM. Influence of the social network on married and unmarried older adults’ mental health. The Gerontologist. 2018;58(6):1109–13.

35. Cwikel J, Gramotnev H, Lee C. Never-married childless women in Australia: Health and social circumstances in older age. Social Science & Medicine. 2006;62(8):1991–2001.

36. Ermer AE, Proulx CM. Associations between social connectedness, emotional well-being, and self-rated health among older adults: Difference by relationship status. Research on aging. 2019;41(4):336–61.

37. Batty GD, Shipley M, Tabák A, Singh-Manoux A, Brunner E, Britton A, et al. Generalizability of occupational cohort study findings. Epidemiology. 2014;25(6):932–3.

38. Sommerlad A, Sabia S, Singh-Manoux A, Lewis G, Livingston G. Association of social contact with dementia and cognition: 28-year follow-up of the Whitehall II cohort study. PLoS medicine. 2019;16(8):e1002862.

39. Holt-Lunstad J, Smith TB, Baker M, Harris T, Stephenson D. Loneliness and social isolation as risk factors for mortality: a meta-analytic review. Perspectives on psychological science. 2015;10(2):227–37.

40. Smith ML, Steinman LE, Casey E. Combatting Social Isolation Among Older Adults in a Time of Physical Distancing: The COVID-19 Social Connectivity Paradox. Frontiers in public health. 2020;8:403.

41. Verhulsdonk S, Quack R, Hoft B, Lange-Asschenfeldt C, Supprian T. Anosognosia and depression in patients with Alzheimer’s dementia. Arch Gerontol Geriatr. 2013;57(3):282–7. Epub 2013/04/20. doi: 10.1016/j.archger.2013.03.012. PubMed PMID: 23597486.

## Citation for Perceived social support questionnaire

Kliem S, Mößle T, Rehbein F, Hellmann DF, Zenger M, Brähler E. A brief form of the Perceived Social Support Questionnaire (F-SozU) was developed, validated, and standardized. Journal of clinical epidemiology. 2015;68(5):551–62.

